# On-scalp magnetoencephalography for childhood epilepsies

**DOI:** 10.1101/2021.09.06.21262839

**Authors:** Odile Feys, Pierre Corvilain, Alec Aeby, Claudine Sculier, Florence Christiaens, Niall Holmes, Matthew Brookes, Serge Goldman, Vincent Wens, Xavier De Tiège

**Author notes:** **Corresponding author:** Dr Odile Feys, Laboratoire de Cartographie fonctionnelle du Cerveau (LCFC), ULB Neuroscience Institute (UNI), Université libre de Bruxelles (ULB), 808 Lennik Street, Brussels, Belgium.

## Abstract

Magnetoencephalography (MEG) is an established method to investigate epilepsy. Current MEG systems house hundreds of cryogenic sensors in a rigid, one-size-fits-all helmet, which results in several limitations, particularly in children. On-scalp MEG based on optically-pumped magnetometers (OPMs) may alleviate these limitations.

We report on five children (5–11 years old) with self-limited focal (n=3) or structural (n=2) epilepsy who underwent cryogenic (102 magnetometers) and on-scalp (32 OPMs) MEG. We compared the two modalities for the detection and localization of interictal epileptiform discharges (IEDs).

We identified IEDs in all children with comparable sensor topographies for both MEG devices. IED amplitudes were 2.3-4.8 times higher with on-scalp MEG and signal-to-noise ratio (SNR) was also 27-60% higher with on-scalp MEG in all but one patient with large head movement artifacts. The neural source of averaged IEDs was located at about 5 mm (n=3) or higher (8.3 mm, n=1; 15.6 mm, n=1) between on-scalp and cryogenic MEG.

Despite limited number of sensors and scalp coverage, on-scalp MEG detects IEDs in epileptic children with higher SNR than cryogenic MEG. This technology, which is in constant development, should become a reference in the diagnostic workup of epilepsy and replace cryogenic MEG in the near future.

## Introduction

Magnetoencephalography (MEG) has proven its clinical added value for the non-invasive localization of the irritative zone in patients with refractory focal epilepsy,^1, 2^ especially for epilepsy originating from outside the temporal lobe that are common in children.^3^ MEG improves patients’ surgical management by (i) detecting irritative zones not captured by conventional EEG, (ii) contributing to subtle brain lesion detection, or (iii) improving intracranial EEG planning/surgical resection accuracy.^2-5^

Current cryogenic MEG systems house hundreds of superconducting quantum interference devices (SQUIDs) in a rigid, one-size-fits-all helmet.^6^ SQUIDs are the main limitation of cryogenic MEG (for details, see ^6^). Because of their need for cryogenic cooling, a thermally insulating gap is required between the scalp and sensor, meaning the brain-to-sensor distance is about 2-5 cm in adults who fit the system well, and even larger in subjects with small heads such as children. Small head size increases the brain-to-sensor signal attenuation (as magnetic fields decrease with the square of the distance) and lead to larger head movements within adult-sized helmets when children struggle to keep still.

Optically-pumped magnetometers (OPMs) are novel cryogen-free magnetic field sensors. They can be placed directly on the scalp to record neuromagnetic signals with an adequate level of noise, even during movements.^6^ Consequently, OPM arrays can adapt to any head shape or size, and record human brain activity in natural conditions.^6^ On-scalp OPM-based MEG (OPM-MEG) should substantially improve signal-to-noise ratio (SNR) and spatial resolution, especially in children.^6^

OPM-MEG recording (15 OPMs, bespoke 3D-printed scanner cast housing the OPMs) has been described in one adult patient with refractory focal epilepsy.^7^ OPM-MEG was able to detect and localize the source of interictal epileptic discharges (IEDs) but a direct comparison with SQUID-MEG was missing.

Here, we study OPM-MEG in the routine clinical use by comparing IED recordings using 32 OPMs fixed on comfortable and flexible EEG-like caps with recordings from SQUID-MEG in five children with extratemporal focal epilepsy.

## Materials and Methods

### Patients

Between April and June 2021, we collected five children with focal epilepsy (4F/1M; median age, 9.4 years; range, 5–11 years; Table 1) who met the inclusion criteria. Three patients had self-limited genetic focal epilepsy, while the two others had refractory focal epilepsy of unknown cause (one with focal hypometabolism on PET with [18F]-fluorodeoxyglucose concordant with the presumed epileptogenic zone (EZ) but without detectable lesion on structural brain 3 Tesla MRI, one who was not seizure-free after a resected right temporal dysembryoplastic neuroepithelial tumor with a distant presumed EZ; Patients 2 and 5 in Table 1). Three of the five patients had epileptic encephalopathy.^8^ Inclusion criteria were (i) self-limited epilepsy with centro-temporal spikes (SL-ECTS) or lesional/non-lesional refractory focal epilepsy, (ii) frequent unifocal IEDs on a previous clinical EEG, and (iii) ability to remain relatively still for at least 15 min of MEG recordings.

**Table 1:**
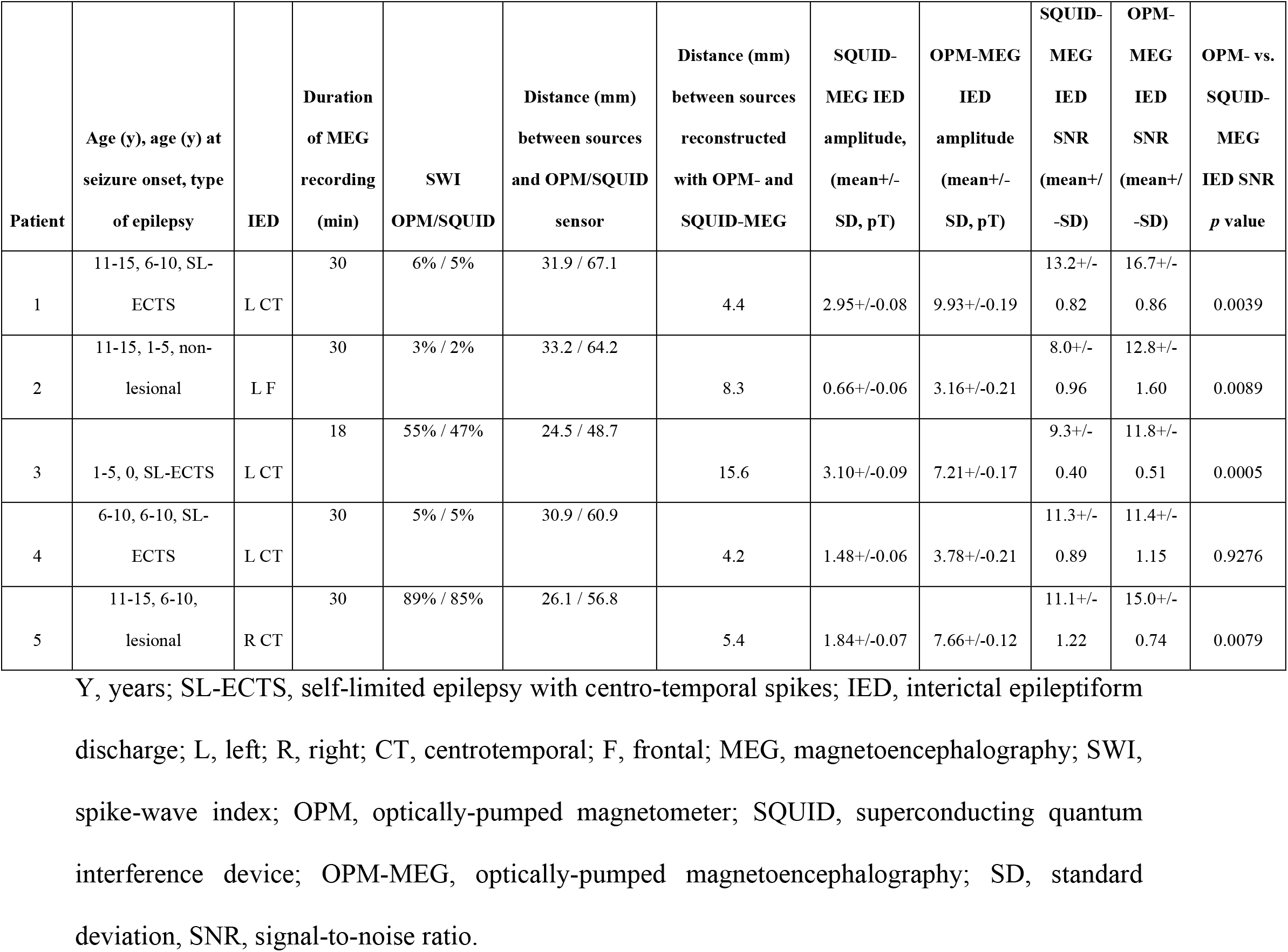
Epileptic children’s clinical characteristics and results of MEG investigations.

| Patient | Age (y), age (y) at seizure onset, type of epilepsy | IED | Duration of MEG recording (min) | SWI OPM/SQUID | Distance (mm) between sources and OPM/SQUID sensor | Distance (mm) between sources reconstructed with OPM- and SQUID-MEG | SQUID-MEG IED amplitude, (mean+/- SD, pT) | OPM-MEG IED amplitude (mean+/- SD, pT) | SQUID-MEG IED SNR (mean+/- SD) | OPM-MEG IED SNR (mean+/- SD) | OPM- vs. SQUID-MEG IED SNR *p* value |
| --- | --- | --- | --- | --- | --- | --- | --- | --- | --- | --- | --- |
| 1 | 11-15, 6-10, SL-ECTS | L CT | 30 | 6% / 5% | 31.9 / 67.1 | 4.4 | 2.95+/-0.08 | 9.93+/-0.19 | 13.2+/-0.82 | 16.7+/-0.86 | 0.0039 |
| 2 | 11-15, 1-5, non-lesional | L F | 30 | 3% / 2% | 33.2 / 64.2 | 8.3 | 0.66+/-0.06 | 3.16+/-0.21 | 8.0+/-0.96 | 12.8+/-1.60 | 0.0089 |
| 3 | 1-5, 0, SL-ECTS | L CT | 18 | 55% / 47% | 24.5 / 48.7 | 15.6 | 3.10+/-0.09 | 7.21+/-0.17 | 9.3+/-0.40 | 11.8+/-0.51 | 0.0005 |
| 4 | 6-10, 6-10, SL-ECTS | L CT | 30 | 5% / 5% | 30.9 / 60.9 | 4.2 | 1.48+/-0.06 | 3.78+/-0.21 | 11.3+/-0.89 | 11.4+/-1.15 | 0.9276 |
| 5 | 11-15, 6-10, lesional | R CT | 30 | 89% / 85% | 26.1 / 56.8 | 5.4 | 1.84+/-0.07 | 7.66+/-0.12 | 11.1+/-1.22 | 15.0+/-0.74 | 0.0079 |

Y, years; SL-ECTS, self-limited epilepsy with centro-temporal spikes; IED, interictal epileptiform discharge; L, left; R, right; CT, centrottemporal; F, frontal; MEG, magnetoencephalography; SWI, spike-wave index; OPM, optically-pumped magnetometer; SQUID, superconducting quantum interference device; OPM-MEG, optically-pumped magnetoencephalography; SD, standard deviation, SNR, signal-to-noise ratio.

This study was approved by the Ethics Committee of the CUB Hôpital Erasme (Reference: P2019/426, B406201941248). Parents and children gave written informed consent prior to their inclusion.

### Data acquisition

OPM-MEG was performed using 32 zero-field magnetometers (Gen-2.0, QuSpin Inc, Colorado, USA; single-axis mode, gain 2.7V/nT) whose signal was fed to a digital acquisition unit (National Instruments, Texas, USA; sampling rate 1200Hz, no band-pass filter). To maximize patients’ collaboration and comfort during prolonged recordings, we adapted the concept of flexible EEG-like cap previously described^9, 10^ for OPM-MEG recordings (Figure 1). We 3D printed plastic sensor mounts (64 per EEG cap, Figure 1, Left) that were sewn on conventional flexible EEG caps (EasyCap, Herrsching, Germany; 2 sizes to adapt to children’s head circumference) according to the 10-10 EEG system to rigidly fix OPMs on the scalp. This design prevented the insertion of any material between the scalp and the cap that represented potential source(s) of discomfort or pain.^9^ The mounts covered about 40% of the inferior part of the OPMs and had vertical openings (Figure 1, Middle) to allow dissipation of OPM-related heat. Each mount was also equipped with one hollow at each corner of the mount base (Figure 1, Middle) to allow quick (about 10 min) and precise digitization of OPM position on children’s scalp using an electromagnetic tracker (Fastrak, Polhemus, Colchester, VT, USA). This flexible cap was easy and quick (1-2min) to install on children’s head. Three small marks were also drawn on the children’s forehead and EEG cap (one right, one middle, one left) using a skin pencil to check that the cap did not move relative to children’s head during acquisition. Sensors were placed around the presumed EZ as determined by previous EEG. Recordings of 15–30 min (according to age) took place inside a compact magnetically shielded room (MSR) optimized for OPM recordings (OPM-compact MuRoom, Magnetic Shielded Limited, Kent, UK), with a background magnetic field of less than 15 nT after deGaussing. Children comfortably sat at the MSR center, watched a movie, with no constraint on head position or movement. Of note, no further field compensation^6^ was applied here. Sensor locations were obtained outside the MSR after the recording and careful removal of OPMs by digitizing the four base points of each mount housing an OPM (Figure 1, Middle) and at least 300 points (face and scalp) relative to anatomical fiducials. This digitization procedure took about 10 min in each child.

**Figure 1.**
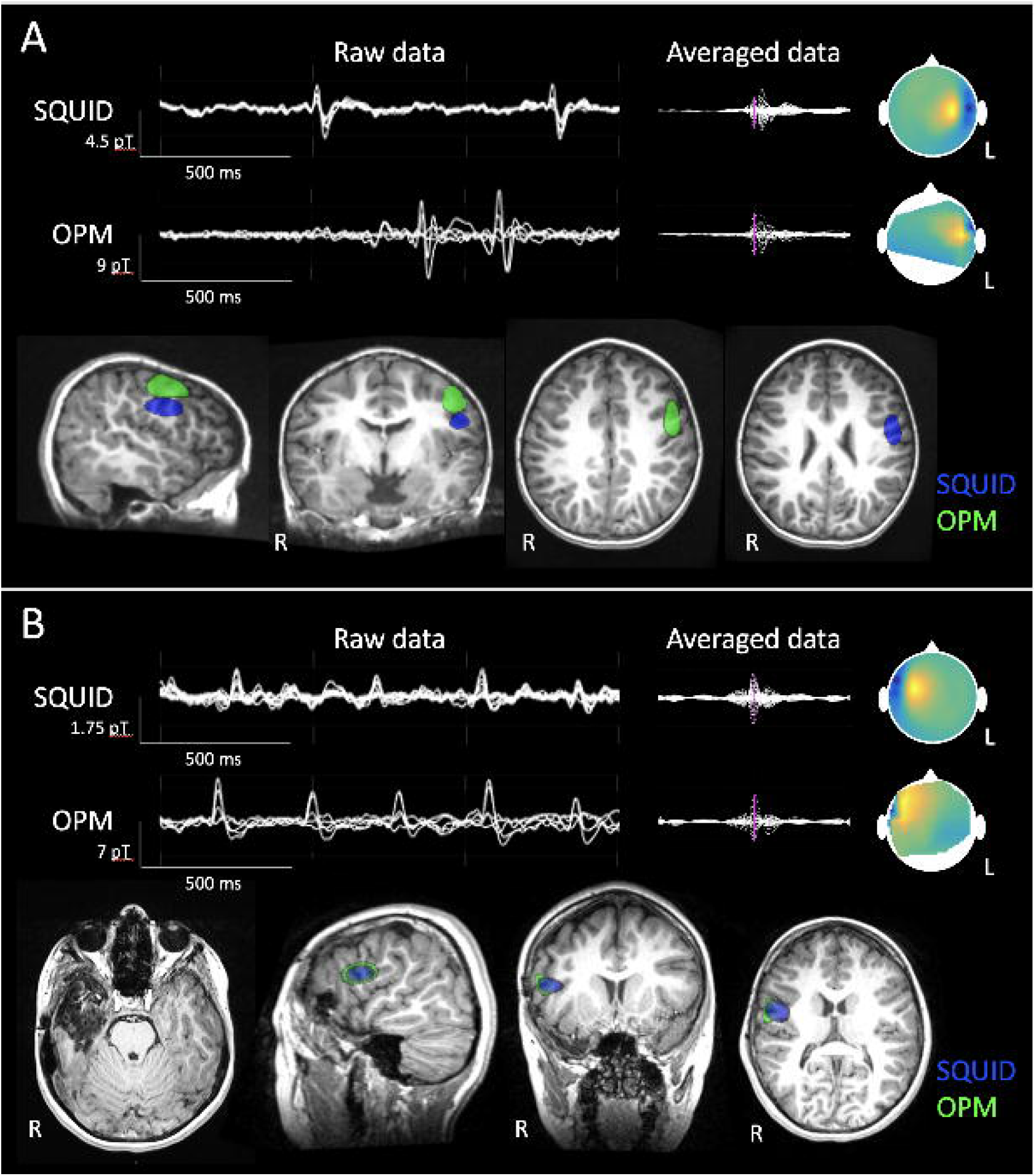
OPM- and SQUID-based MEG. **Left**. Illustration of the MSR dedicated to OPMs. **Middle left**. Illustration of the flexible EEG-like cap set-up used in this study for OPM-MEG recordings. **Middle right**. Illustration of the 3D printed plastic mount sewn on the EEG cap and housing one OPM oriented radially. Mounts covered about 40% of the inferior part of the OPMs and had vertical openings to allow dissipation of OPM-related heat. Each mount was also equipped with one hollow at each corner of the mount base (small rounds with hollow) to allow quick and precise digitization of OPM position on children’s scalp. **Right**. Illustration of the SQUID-MEG used in this study.

SQUID-MEG (Figure 1, Left) was recorded in similar conditions using a commercial 306-channel, whole-scalp neuromagnetometer placed in a light-weight MSR (Triux and Maxshield, MEGIN, Helsinki, Finland; 204 planar gradiometers, 102 magnetometers; sampling rate 1000Hz, band-pass filter 0.1-300Hz).^11^ Head movements were tracked by four position indicator coils. These coils and 300 face/scalp points were digitized relative to anatomical fiducials using the same electromagnetic tracker.

OPM-MEG was done before SQUID-MEG in all children.

Each patient also underwent a 3D T1-weighted brain MRI, either during their clinical assessment (1.5 T Intera, Philips, The Netherlands) or after the MEG (3T Signa PET-MR, GE Healthcare, Illinoi, USA).

### Data preprocessing

Both MEG data were denoised using distinct spatial filters, i.e., principal components analysis for OPMs (3 first components associated with slow, large-amplitude drifts and movement artefacts were removed) and signal space separation with movement correction (Maxfilter, MEGIN) for SQUIDs.^12^ Signals were then band-pass filtered at 3–40 Hz for IED detection. For comparability with OPM-MEG, SQUID-MEG was restricted to its 102 magnetometers. For source reconstruction, the MRI was manually co-registered to OPM-MEG and SQUID-MEG separately using their respective digitalization (MRIlab, MEGIN). Forward models were computed for both modalities using the one-layer boundary element method (MNE-C^13^) based on MRI tissue segmentation (Freesurfer^14^). For OPM-MEG, sensor locations and orientations were estimated from the digitalization.

### Data analysis

IEDs were visually identified in MEG signals by trained neurologists (O.F., X.D.T.). A spike-wave index (SWI) was computed for each data set (i.e., OPM- and SQUID-MEG) and patient as in ^15^. Data were epoched from -300 to 300 ms after each spike event, baseline corrected (from -100 ms to -50 ms) and finally averaged. The neural source at the peak of averaged spikes were localized using an home-made implementation of dynamic statistical parametric mapping^16^ (noise covariance estimated from baseline data, regularization from the global SNR^17^).

Peak amplitude and SNR of IEDs were estimated at each spike event for the sensor showing maximum averaged spike amplitude and compared across modalities using two-sided unpaired *t* tests. This allowed comparing two unequal sets (OPM-vs. SQUID-MEG) of IEDs for each patient. Finally, the distance between the reconstructed neural sources and the closest OPM or SQUID sensor was estimated to assess how much closer OPM sensors were from the brain compared with SQUIDs.

### Data availability

Data are available upon reasonable request to the corresponding author and after approval of institutional (i.e., CUB Hôpital Erasme & Université libre de Bruxelles) authorities.

## Results

Table 1 summarizes the results of all children. Figure 2 illustrates the data obtained in patients 3 and 5.

**Figure 2.**
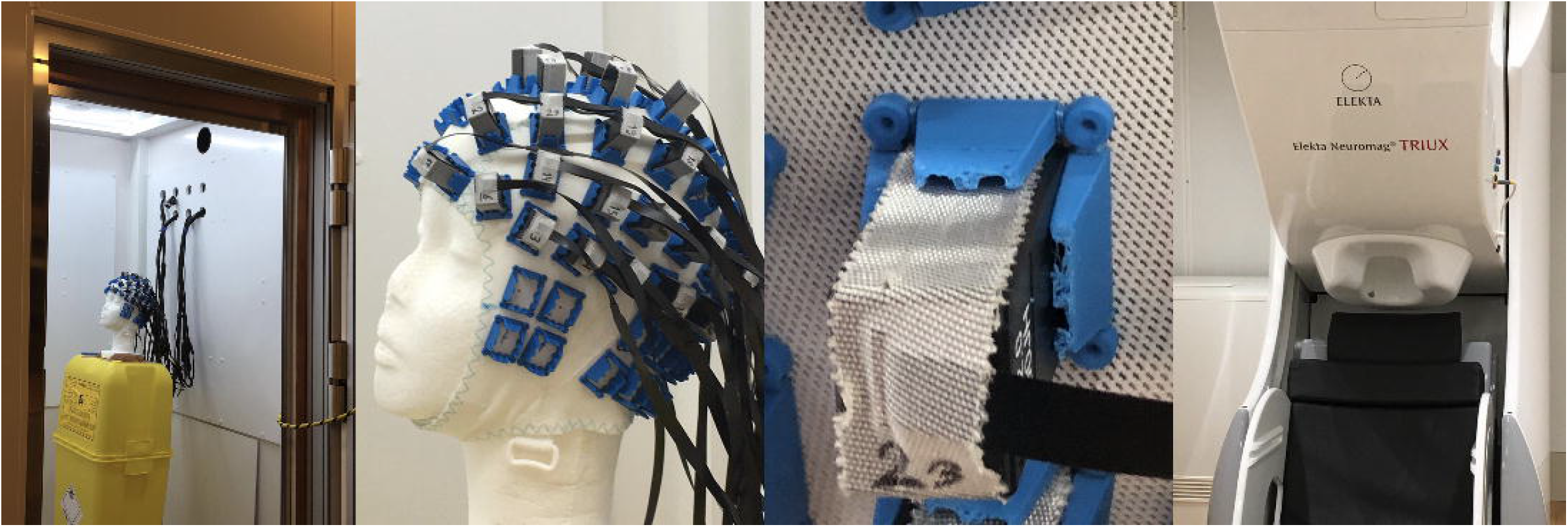
SQUID- and OPM-MEG data. **(A)** Patient 3. **Top, Left**. Sample of the background brain activity and of some IEDs recorded with SQUID- and OPM-MEG. Signals from a selected group of magnetometers were superimposed. **Top, Right**. Averaged IED signals along with the magnetic field topography (sensor array viewed from top, arbitrary scale) at the spike peak (indicated by purple vertical line). **Bottom**. Neural source reconstructions obtained at the averaged IED peak and displayed on parasagittal (Left; left hemisphere), coronal (Middle) and two axial (Right) brain MRI slices. The distance between locations of maximum source activity for OPM- and SQUID-MEG was 15.6 mm. **(B)**. Patient 5. Legend is similar to A. **Bottom**. Axial brain MRI slice (Left) illustrating the resection cavity of the right temporal dysembryoplastic neuroepithelial tumor. Source reconstructions with OPM- and SQUID-MEG are displayed on parasagittal (Left; right hemisphere), coronal (Middle), and axial (Right) slices. The localization difference was 5.4 mm.

Both types of MEG recordings were well tolerated by the children and resulted in good data quality in all but one child (Patient 4), who exhibited episodic large movement artefacts in OPM-MEG data. No misalignment of the EEG cap relative to children’s forehead was noticed based on the three marks.

Unifocal and monomorphic IEDs were found in all children with SWI ranging from 2 to 89%. At the sensor level, IEDs had comparable sensor topographies for both types of MEG and were consistent with previous clinical EEG. IEDs amplitude was systematically higher (from 2.3 to 4.8 times higher) with OPM-MEG compared with SQUID-MEG, with high statistical significance (negligibly small *p* value). Their SNR was also significantly higher (from 27 to 60% higher) with OPM-MEG in all but one (Patient 4).

At the source level, the distance of the neural source of the averaged IEDs peak ranged from 4.2 mm to 15.6 mm between OPM- and SQUID-MEG reconstructed sources. The mean distance between the reconstructed neural source and the closest sensor was 29.4 mm for OPM-MEG and 57.6 mm for SQUID-MEG.

## Discussion

This case series demonstrates that, compared with SQUID-MEG, multichannel OPM-MEG (i) is well tolerated in five children aged 5-11 years with focal epilepsy, (ii) accurately detects IEDs, (iii) provides significantly higher IED amplitude in all children, (iv) provides significantly higher IED SNR in all but one child, and (v) locates similar or close neural sources of IEDs. This was achieved despite a smaller number of sensors (32 vs. 102) and consequent limited scalp coverage.

To fit routine clinical use in epilepsy, we adapted the flexible EEG cap^9, 10^ for on-scalp OPM recordings to maximize patients’ collaboration and comfort. This light and flexible cap design was operator- and child-friendly, and contributed to the excellent tolerance of OPM-MEG by the epileptic children, already used to undergoing EEG investigations. It also has the advantage of placing sensors at the scalp surface, which is difficult to reach with rigid helmets. One possible disadvantage is that any movement of the OPM relative to the scalp during the recording will lead to artifacts and inaccuracies in the source reconstruction. The mount design also facilitated the OPM localization procedure that took about 10 min in each child, which was well tolerated and reasonable. In the near future, the adaptation of optical co-registration techniques^8^ will certainly allow to speed up and increase the accuracy of this localization.

IEDs were detected in all patients with comparable SWI between OPM- and SQUID-MEG. IED peak amplitudes were 2.3 to 4.8 times higher with OPM-than with SQUID-MEG. This merely reflects the reduced (∼ 3 cm on average) brain-to-sensor distance afforded using OPMs.^6^ Still, OPM signals were generally noisier than SQUID signals, although intrinsic sensor noise is similar^18^, because children were free to move and consequent OPM movements created signal artefacts commensurate to the background magnetic environment. By opposition, SQUIDs are fixed and subjected to efficient software denoising.^12^ Importantly, despite these disadvantages, IED peak SNR remained significantly higher with OPM-MEG in four children and similar than SQUID-MEG in the other with large movements (Patient 4), which suggest that our EEG cap setup is adequate for pediatric recordings of IEDs. Movement-related artefacts in OPM signals could be reduced substantially with extra hardware solutions such as field nulling coils^6^ (which would likely reduce background fields from ∼15 nT to < 1 nT, and consequently reduce movement artifacts by a similar factor) and the development of OPM denoising algorithms.^19, 20^ Therefore, the quality of OPM-MEG recordings in epileptic children will continue to improve in the coming years to fully overpass SQUID-MEG.

Differences in the location of IED reconstructed neural sources based on OPM- and SQUID-MEG signals were in the range of SQUID-MEG spatial resolution (i.e., about 5 mm; 3 patients) or higher (2 patients). The latter could be related to different IED neural generators (non-simultaneous recordings), differences in the number and spatial coverage of sensors, inaccuracies in the digitization procedure, or the higher SNR of OPM signals. Still, OPM-MEG based on 32 sensors placed around the presumed EZ can identify similar IED neural generators than SQUID-MEG. An increase in the number of sensors for OPM-MEG and the development of triaxial OPM sensors^20^ should position OPM-MEG as the future reference for clinical MEG investigations.

This study was limited by the difficulty to compare the sensitivity of IED detection of both modalities intrinsically associated with non-simultaneous recordings, the absence of a reference standard (e.g., intracranial recording, resection cavity) to compare the spatial precision of source reconstructions (see ^21^ for a discussion on this issue), the limited number and spatial coverage of OPMs, and the small number of children investigated. Still, it provides unprecedented evidence supporting the clinical added value of OPM-MEG compared with SQUID-MEG. OPM-MEG have major advantages of in the field of epilepsy. Compared to EEG, they are easy to use with no need of electrolyte gel or electrode gluing on the scalp. Compared to SQUID-MEG, there is no need of head position indicator coils, they are movement-friendly, which opens the possibility of routine ictal MEG and prolonged video-MEG recordings, and their cost is reduced. This nascent technology has therefore all the potential to supplant SQUID-MEG in the clinical management of epileptic patients. As a future method of reference, it is expected to even replace scalp EEG in certain circumstances.

## Data Availability

Data are available upon reasonable request to the corresponding author and after approval of institutional (i.e., CUB Hopital Erasme & Universite libre de Bruxelles) authorities.

## Abbreviations

EZ: Epileptogenic zone
IED: Interictal epileptiform discharge
MEG: Magnetoencephalography
MSR: Magnetic shielded room
OPM: Optically-pumped magnetometer
OPM-MEG: Optically-pumped magnetometer based magnetoencephalography
SL-ECTS: Self-limited epilepsy with centro-temporal spikes
SNR: Signal-to-noise ratio
SQUID: Superconducting quantum interference device
SWI: Spike-wave index

